# Studies on Geographic Variations and Gender Bias in Thyroid Cancer at National Guard Hospitals in Saudi Arabia

**DOI:** 10.1101/2024.01.26.24301836

**Authors:** Nawaf Alanazi, Zainab Ibrahim Alabbad, Nahal Hamad Almutairi, Rahmah Abdulwahab Alhulaymi, Jawaher Raed Alfalah, Eithar Qasim Alkhalifa, Rehab Ibrahim Abubaker, Kanza Adeel, Sarah Balghonaim, Alzahra Alshayeb, Sarah Al-Mukhaylid, Zafar Iqbal

## Abstract

Thyroid cancer (TC) is one of the most prevalent cancers in the world, ranked as ninth worldwide and third in Saudi Arabia. This report reviews the incidence of TC at National Guard Hospitals (Riyadh, Jeddah, and Ahsa), in relation to gender, geographic variation and age. The study design is quantitative cross-sectional retrospective review; data of 2166 patients with TC diagnosed in National Guard Hospitals (Ahsa -Jeddah- Riyadh) in the period of 2015-2021 was received from King Abdullah International Medical Research Center (KAIMRC). Chi- square, t-test, and ANOVA-tests were used in analyzing the relationship between variables. Out of the 2166 patient records, (79.78%) were females, and (46.6%) were in the age group of 40-60 with a mean age of 47.28 years. Riyadh had the highest incidence of TC (49.8%), while Ahsa had the lowest (5.6%). the commonest TC type was papillary (83.5%) followed by follicular (4.0%), medullary (1.2%), anaplastic (1.1%), hurthle cell carcinoma (0.8%) and thyroid lymphoma (0.1%). Females were more suspected to have TC, and the risk of having TC increases as age increases until the age of 60, where it decreases. The registered cases were more in Riyadh, Jeddah and Ahsa, respectively, except for lymphoma and hurthle cell, as they were high in Jeddah. Lymphoma was reported in Jeddah and in males only. The mortality rate was low (2.4%). However, increased death risk was observed in patients diagnosed above the age of 60. Mortality was seen in papillary, anaplastic and a single death case was reported in follicular TC.

**Research highlights:**

- Females are at higher risk to develop thyroid cancer than males.
- The risk of having thyroid cancer increased by age in both genders until the age of 60 where it decreased.
- The registered cases were higher in Riyadh then Jeddah, and it was the lowest in Ahsa.
- Lymphoma and hurtle cell were higher in Jeddah.
- Mortality was low and seen in papillary, anaplastic and a single death case was reported in follicular thyroid cancer.

## Introduction

The thyroid gland is one of the endocrine glands located at the base of the neck. It produces thyroxine (T4) and triiodothyronine (T3) hormones which are important for the growth and regulation of vital body functions such as breathing and blood pressure [1]. One of the series of disorders that affect the thyroid gland is cancer, which is an uncontrolled growth of thyroid cells resulting in a mass called a tumor in combination with irregular production of hormones. It is categorized into various types based on the affected cell type [2]. The commonest form of thyroid cancer is papillary thyroid cancer, and it is the least serious due to the slow growth of the cancerous cells [3]. Secondly, follicular thyroid cancer is thought to be related to iodine deficiency [2]. Thirdly, Hurthle cell cancer is classified in some articles as a subtype of follicular thyroid cancer, but it grows more aggressively [4]. Fourthly, anaplastic thyroid cancer is the most aggressive one due to the ability of the malignant cells to metastasize fast affecting the lymph nodes around the thyroid gland in the early stage of the disease [5]. Fifthly, medullary thyroid cancer is commonly associated with multiple endocrine neoplasias [6]. Lastly, Thyroid lymphoma is a rare malignancy in which thyroid lymphocytes transform into malignant cells [7]. Thyroid cancer is asymptomatic earlier in the disease. Though when it grows, it can cause a palpable lump (nodule), and changes in a patient voice (including hoarseness and difficulty swallowing) [8].

The general laboratory tests for thyroid function such as thyroid-stimulating hormone (TSH), T4, and T3 levels tests are performed to diagnose thyroid cancer [9–10]. Follow-up tests are required to differentiate between different types of thyroid cancer such as nuclear medicine imaging studies and microscopic fine-needle aspiration (FNA) and core biopsy [8,5]. The etiology of thyroid cancer is still unclear, but some factors have been associated with increasing the risk [11]. One of the greatest risk factors is radiation exposure. Studies show a relationship between the age of exposure and the risk of developing thyroid cancer. It states that the younger the age is during the exposure, the greater the risk of developing thyroid cancer [12]. Another risk factor is the history of having a benign thyroid condition such as thyroid nodule, goiter, and thyroiditis [13]. In addition, individuals with a family history of thyroid cancer are at higher risk than others to develop one. Also, hereditary conditions such as multiple endocrine neoplasia and familial adenomatous polyposis (FAP) are known to be associated with thyroid cancer, but these are rare conditions [11]. Furthermore, acromegaly is a rare pituitary tumor leading to excessive growth hormone secretion causing different organs, including the thyroid, to grow which increases the risk of thyroid cancer [14]. Also, diet can increase the risk, especially a low-iodine diet. Moreover, hormonal factors can play a role in the females’ reproductive years [11].

Thyroid cancer is one of the most common types of endocrine cancer around the world [15]. According to a global study done by the American Cancer Society in 2020, the number of new thyroid cancer cases was 586,202. The males represent 23.42% of the cases, while the females represent 76.58%. As a result, the incidence of cancer among females was three times higher than among males. Moreover, the mortality cases of thyroid cancer in 2020 was 43,646 which represents approximately 7.45% of all the cases [16].

In the Middle East, there is a noticeable increase in the incidence of all cancer forms. Projections show that by 2030, the cases will increase approximately 1.8 times [17]. Saudi Arabia has the highest rate of thyroid cancer among other middle eastern countries, with age-standardization rates (ASR) of 7.0. Next, Lebanon with an ASR of 6.0, and then Morocco with an ASR of 5.8. In the Arab Gulf states, Kuwait comes second, with an ASR of 5.2 then the UAE with an ASR of 5.0 [18]. A recent study conducted in Saudi Arabia showed that thyroid cancer incidence rates ranked third after breast and colorectal cancer as the most common cancers [19,20]. The number of new cases registered in 2020 was about 2,833 [19]. The capital, Riyadh, ranked first in the number of registered cases. The median age of diagnosis was 39 years in females and 44 years in males [20]. Females are more susceptible to developing thyroid cancer. Approximately, 13.9 new cases of thyroid cancer were diagnosed for every 100,000 females, while the incidence in males was only 4 out of 100,000 males. The mortality rate of thyroid cancer was low compared to the incidence rate, about 1 per 100,000 [18].

Thyroid cancer is one of the commonest cancers globally and in Saudi Arabia, especially among females. However, limited studies have been conducted to assess the geographical variations and gender bias on thyroid cancer within Saudi Arabia’s regions. Therefore, this study aims to evaluate the geographical variations of thyroid cancer among Al Ahsa, Jeddah, and Riyadh cities and gender bias at National Guard Hospitals (NGHA).

## Materials and methods

The study was performed at King Saud bin Abdulaziz University for Health Science in collaboration with the Oncology Medicine department of the National Guard Hospitals (Riyadh, Jeddah, Alahsa) from 2021 to 2022. The study population includes all patients diagnosed with thyroid cancer in the three hospitals from 2015 to 2021. The study design was a quantitative, cross-sectional, retrospective chart review.

Data of confirmed patients with thyroid cancer from Al-Ahsa, Jeddah, and Riyadh National Guard Hospitals (NGHA) were obtained from medical records at the King Abdullah International Medical Center (KAIMRC).

The received data included 2271 records of thyroid cancer patients. 105 records were excluded because they were incomplete, repeated, or from cities other than Jeddah, Riyadh, or Ahsa. 2166 records were included in the study, and 788 of them included data about the patient history of diabetes, hypertension, and hyperlipidemia. Then, the data was analyzed in Social Sciences Software (SPSS). Categorical data were analyzed and reported as proportion, while quantitative data were reported as standard deviation using chi-square or t-test.

## Results

The total number of thyroid cancer cases in Riyadh, Jeddah, and Ahsa at the National Guard Hospitals was 2166 between the year 2015-2021. Out of 2166 cases, 1728 (79.78%) were females, and 438 (20.22%) were males (Table1). 2046 (94.46%) of the cases were Saudi, and 120 (5.54%) were non-Saudi (Figure 1).

**Figure 1:**
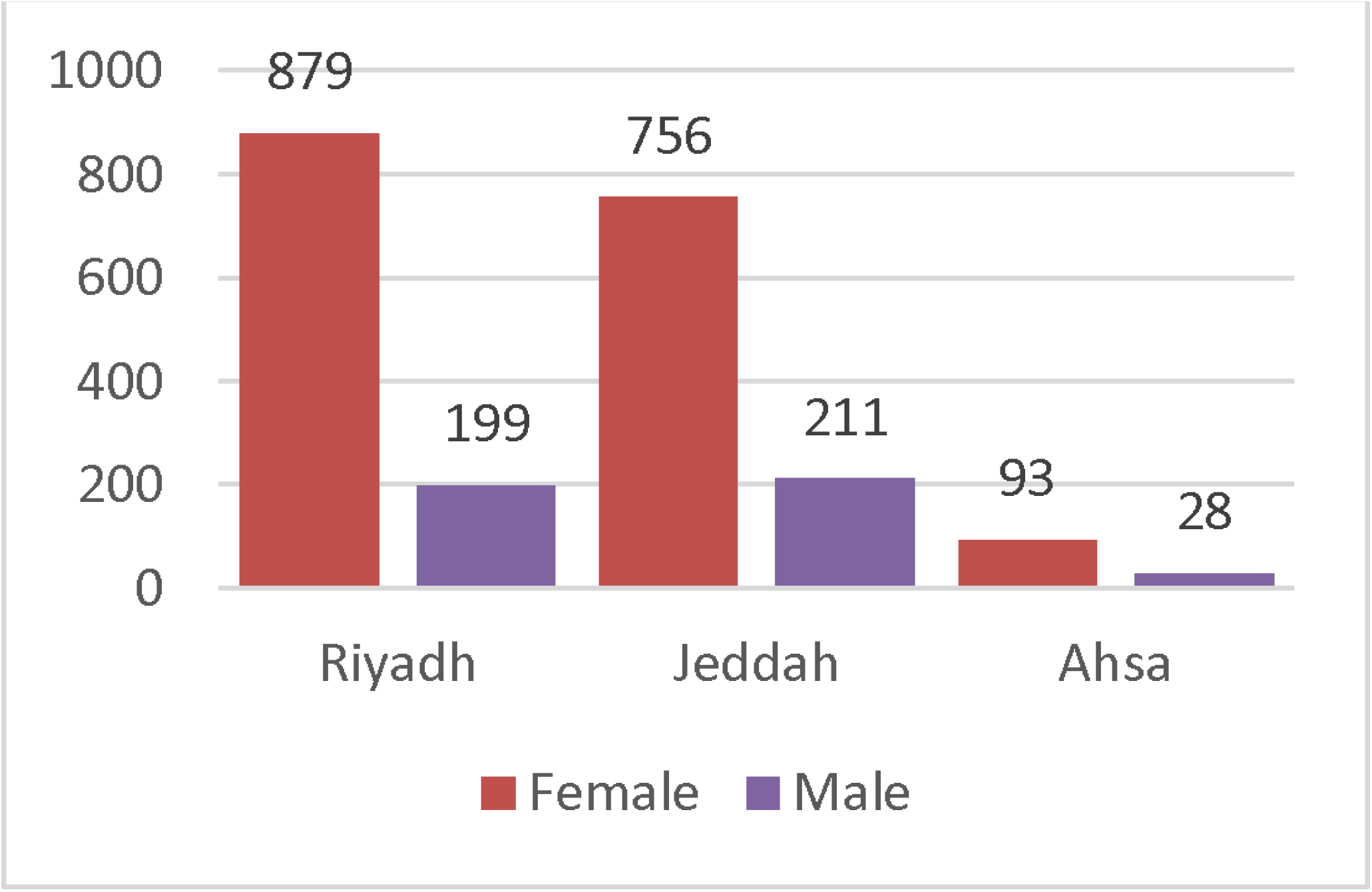
Nationality of Thyroid Cancer Patients

Region-wise, 1078 (49.77%) cases were from Riyadh, 967 (44.64%) cases were from Jeddah, and 121 (5.59%) cases were from Ahsa (Table 1). The patients were divided into four age groups: <20, 20-40, 40-60, and >60, and the number of new cases was 48 (2.2%), 681 (31.4%), 1009 (46.6%), and 428 (19.8%), respectively (Table 1).

**Table 1:**
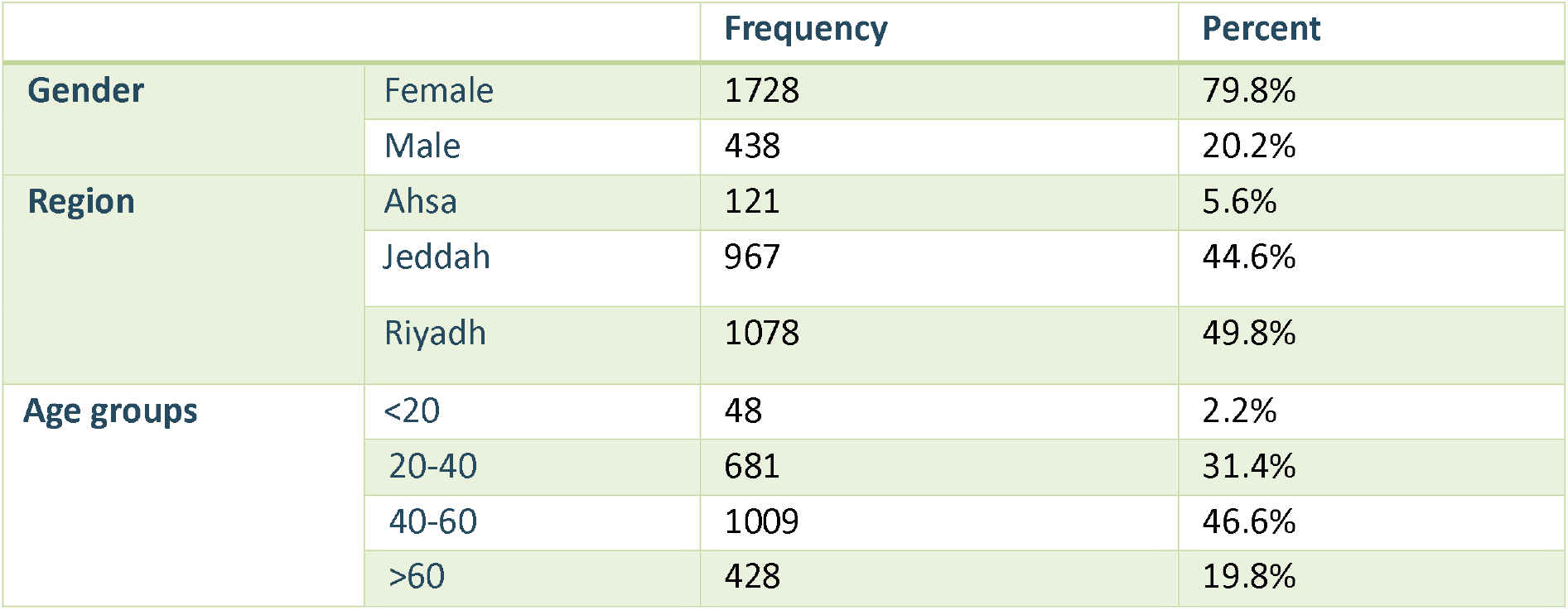
Frequency of Thyroid Cancer Depending on Gender, Region, and Age Group.

|  |  | Frequency | Percent |
| --- | --- | --- | --- |
| Gender | Female | 1728 | 79.8% |
|  | Male | 438 | 20.2% |
| Region | Ahsa | 121 | 5.6% |
|  | Jeddah | 967 | 44.6% |
|  | Riyadh | 1078 | 49.8% |
| Age groups | <20 | 48 | 2.2% |
|  | 20-40 | 681 | 31.4% |
|  | 40-60 | 1009 | 46.6% |
|  | >60 | 428 | 19.8% |

In 2016, the number of cases increased dramatically reaching 567 (26.2%) cases. However, the number of new thyroid cancer cases has decreased over the last four years (Figure 2).

**Figure 2:** Thyroid Cancer Cases in Saudi Arabia from 2015 To 2021

The number of thyroid cancer cases was variable in different geographical areas. In 2015-2021, Riyadh had the highest number of new cases accounting for 1078 (49.8%), out of which 879 (81.53%) of the cases were females and 199 (18.64%) were males with a female-to-male ratio of 4.42:1. Whereas in Jeddah, the total number of cases in the same period was 967(44.6%), 756 (78.17%) of the cases were females and 211 (21.82%) were males with a ratio of 3.84:1. Although the total number of cases in Jeddah was less than in Riyadh, the number of male cases was higher in Jeddah. In Ahsa, there were 121 (5.6%) cases including 93 (76.85%) females and 28 (23.14%) males with a ratio of 3.32:1 (Figure 3).When the ages were compared, the mean age at diagnosis was 47.28 (SD=15.08). It was 51.96 (SD=16.39) years for males and 46.09 (SD=14.5, p-value<0.001) years for females.

**Figure 3:** The Incidence of Thyroid Cancer in Females and Males (Riyadh, Jeddah, Ahsa)

**Figure 4:** The Mean Age of Thyroid Cancer Patient at Diagnosis

Furthermore, the mean age at diagnosis in different geographical areas of the patients was 48.52 years in Riyadh, 45.83 years in Jeddah, and 47.86 years in Ahsa. Moreover, the mean age at diagnosis in each of the three areas for females was 47.43 (SD=14.387), 44.56 (SD=14.427), 45.88 years (SD=15.175) in Riyadh, Jeddah, and Ahsa, respectively, and for males was 53.33 (SD=16.23), 50.34 (SD=16.880), 54.43 years (SD=12.553) in Riyadh, Jeddah, and Ahsa, respectively (Figure4). In addition, to see if there was any difference in the mean age of the three areas, a one-way ANOVA test has been done. The p-value was <.001 (<.05) which means a significant difference between the mean age at diagnosis in the three areas. However, when the mean age of each gender in three areas was compared using two-way ANOVA, the p-value was 0.712 which is more than 0.05 indicating that there was no significant difference between both genders’ means of age among the three areas.

788 of the reported thyroid cancer patients have been diagnosed with other medical conditions, which include: 511 diabetes cases, 421 hyperlipidemia cases, and 522 hypertension (HTN) cases. Out of the 788 patients, 136 (17.26%) were only diabetic, 120 (15.23%) only had hyperlipidemia, 87 (11.04%) only had hypertension, 43 (5.46%) had diabetes and hypertension, 111 (14.08%) had diabetes and hyperlipidemia, 70 (8.88%) had hypertension and hyperlipidemia, while the number of patients who had the three diseases was 221 (28.05%) (Figure 5).

**Figure 5:** Frequency of Hypertension, Hyperlipidemia, and Diabetes among Thyroid Cancer Patients

Data demonstrated that there was a variation in the incidence of thyroid cancer according to age group and gender. In those who were younger than 20, most of the cases were females 42 (87.5%), while males were only 6 (12.5%). With the increase of age, there was an increase in the incidence in males as follows, 104 (15.3%) in the age group 20-40, and 189 (18.7%) in the 40-60 age group, but the number of cases decreased in the patients who are above 60 years 139 (32.5%). Also, females’ cases increased with age. 577 (84.7%) of female cases were in the 20-40 age group, 820 (81.3%) cases were in the 40-60 age group, and, as in males, cases decreased in female patients who are above 60 years 289 (67.5%) (Table2). Using the chi-square test, it can be indicated that the greatest incidence of thyroid cancer in both genders is in the age group of 40-60 years old (p-value <0.001). Therefore, there is strong evidence to reject the null hypothesis and conclude that there is a significant association between gender and age group in patients with Thyroid Cancer (Table 2). When the types of thyroid cancer were assessed, it was found that there were 1809 (83.5%) cases of papillary thyroid cancer followed by follicular thyroid cancer 86 (4.0%), medullary thyroid cancer 25 (1.2%), anaplastic thyroid cancer with 23 (1.1%), Hurthle cell thyroid carcinoma 17 (0.8%), and the last type was lymphoma with 2 (0.1%) cases. In medullary thyroid cancer, the number of incidences between males and females was close with one case difference. In lymphoma, there was a single case reported in a male. On the other hand, the incidence of anaplastic thyroid cancer, follicular thyroid cancer, Hurthle cell thyroid carcinoma, and papillary thyroid cancer was higher in females (Figure 6).

**Table 2:**
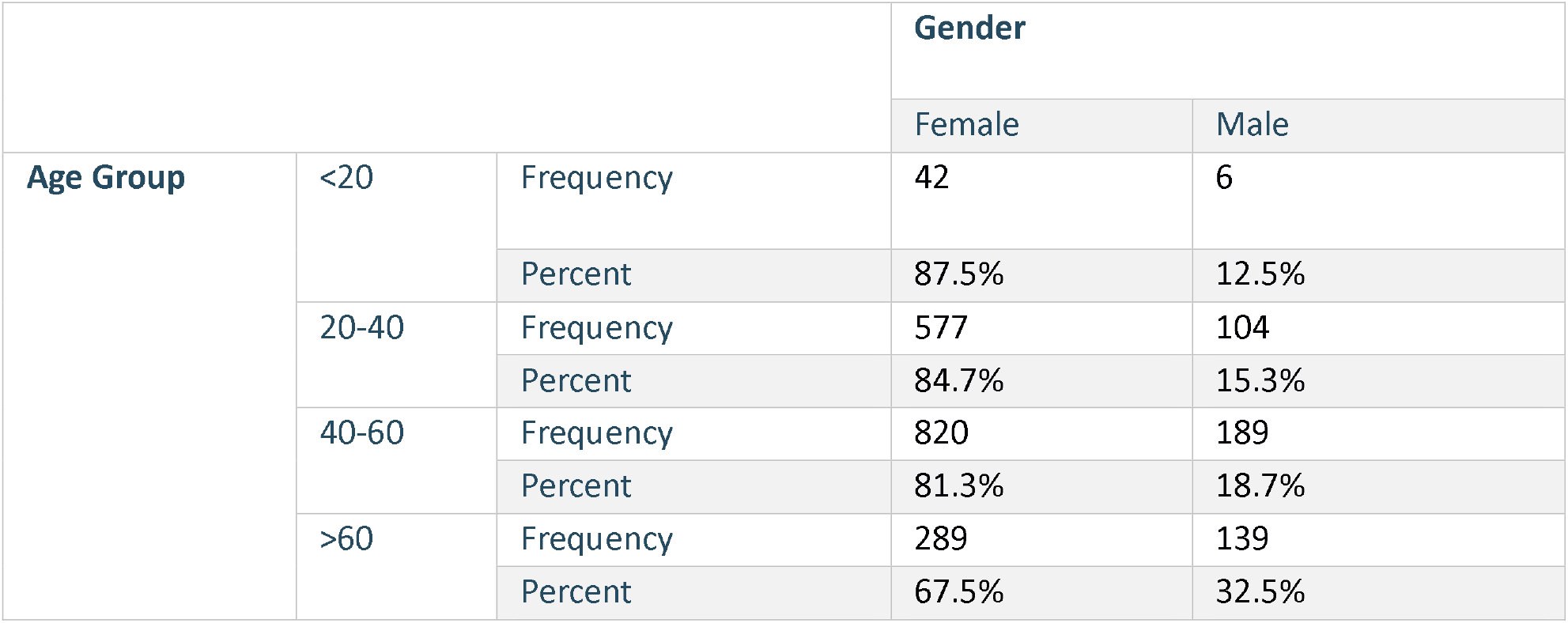
Age Group and Gender.

**Figure 6:** Frequency of Thyroid Cancer Types among Genders

204 (9.4%) cases were not type specified. For the above analysis using the Chi-square test, the p-value was less than <.001 (<.05), so there is an association between gender and thyroid cancer types. The incidence of all thyroid cancer types was higher in Riyadh, except for hurthle thyroid cancer and lymphoma cancer where both were higher in Jeddah. Lymphoma was noticed only in Jeddah.

There were 53 (2.4%) death cases. The mortality was higher in males with 17 cases (3.89%) than in females with 36 cases (2.08%) (Figure 7). Moreover, the mortality in both genders increased as the age of diagnosis increased. Patients diagnosed at ages over 60, had the highest mortality cases accounting for 41 (77.4%) cases out of 53 death cases (Figure 8). When comparing the mortality rate among Al Ahsa, Jeddah, and Riyadh cities, Riyadh had the highest number of deaths with 37 (3.4%) cases, Al Ahsa with 2 (1.7%) cases, then Jeddah had the lowest mortality rate with 14 (1.7%) cases. For the mortality among thyroid cancer types, reported deaths in papillary were 37, anaplastic 15, and a single death case was reported in follicular thyroid cancer. Out of the 23 anaplastic thyroid cancer cases, 15 patients died which represents 65% of the anaplastic cases.

**Figure 7:** Mortality between Males and Females

**Figure 8:** Mortality and Age Group

## Discussion

From the analysis above, key findings show that females had the greatest number of incidences among all age groups and geographic areas with a female-to-male ratio of 4:1, which is constant with the previously reported incidence [22]. Whereas United Arab Emirates’ female-to-male ratio was as low as 2.4:1 [20–21]. The higher susceptibility of females to thyroid cancer is still to be investigated, but a possible cause of this finding refers to the presence of estrogen hormone which works as a stimulating hormone for cancerous cells. Also, the analysis showed that 94% of diagnosed cases are of Saudi citizens, and that was expected as the National Guard hospitals are governmental hospitals that serve mainly Saudi citizens.

The mean age at diagnosis in the recent study was 47.28 ± 15 which is constant with what was reported to be as low as 40.6 and 45.2 in other Saudi Arabian studies [22–23] and 42 ± 14 in the United Arab Emirates study [21]. Moreover, the age group with the highest incidence was 40-60. After the age of 60, there is a decrease in the incidence which is comparable to the Saudi cancer incidence report published in 2015 [20]. However, another study was conducted in Saudi Arabia in 2013 where the highest age group was 30- 39 from 2000 to 2010 [23]. Similarly, in another study in the United Arab Emirates, the highest incidence was in patients younger than 45 [21]. In addition, a study done at First Hospital of China Medical University showed that the highest number of reported cases were for patients older than 60 [24].

Furthermore, the regional distribution showed that Riyadh ranked first in the number of cases, then Jeddah, and the least was Ahsa. This finding is compatible with the Saudi cancer incidence report [20]. That could be due to the number of patients in each hospital as the Riyadh hospital is larger and prepared to treat a higher number of cases. Furthermore, the population is a focal point to consider. All the thyroid cancer types were higher in Riyadh, followed by Jeddah, and lowest in Ahsa except for lymphoma and Hurthel cell cancer, as both were higher in Jeddah. The possible reason for high cases in Riyadh and Jeddah as compared to Ahsa is due to the high population and development of the Riyadh hospital as the main hospital.

The ranking of thyroid cancer types was as follows: papillary, follicular, medullary, anaplastic, Hurthle, and lymphoma ranked last. All of them had more female-reported cases than males except for lymphoma and medullary. A similar increase in male cases of lymphoma as compared to females’ has also been reported in a study conducted at King Fahd Armed Forces in Jeddah [23]. Few articles attributed this to estrogen’s direct impact on lymphoma cell growth, or its effect on the anti-tumor immune response [25].

Through the years 2015-2021, the number of new thyroid cancer cases peaked in 2016. Then, it decreased. Similarly, an article describing the trend of cases in the United States justified the peak of cases in 2007-2017 to be influenced by the new and highly sensitive diagnostic tests leading to increased detection of cancers. Moreover, they found that the number of diagnosed cases decreased afterward [26]. Nonetheless, this study’s 2015 data could not be compared to 2016 data, since the 2015 data started from August and not from January.

In this study, the mortality percentage was 2.4%. A high number of death cases occurred in patients older than 60 years. As for the mortality among thyroid cancer types, 15 (65%) out of the 23 anaplastic thyroid cancer patients died, due to the difficulty of treating this type [5].

## Conclusion

The results showed that females were at higher risk to develop thyroid cancer than males, and the risk of having thyroid cancer increased by age in both genders until the age of 60 when it decreased. The registered cases were higher in Riyadh then in Jeddah, and it was the lowest in Al Ahsa for all thyroid cancer types, except for lymphoma and hurtle cell, which were higher in Jeddah. Lymphoma was reported in Jeddah and males only. Mortality was low, and it was only seen in papillary and anaplastic thyroid cancer, and a single death case was reported in follicular thyroid cancer. Most death cases were noticed in patients older than 60 with a death rate of 1 out of 10 patients.

Some limitations should be noted. First, the data was from the National Guard Hospital only, therefore other patients with thyroid cancer in the same areas were not included in the study. Moreover, there was missing information on the thyroid cancer types and underlying diseases. Besides that, the data started from August 2015 and not from January 2015 (the beginning of the year) to September 2021, not to December 2021 (end of the year) so the data for 2015 and 2021 were incomplete.

It is recommended to include data from other hospitals in the same area. Also, more research is needed on the risk factors in each region as well as the cause of the increase in incidence in 2016.

## Supporting information

https://1drv.ms/f/s!Ai2Oaa5jGDDxgZQtKjFKpvWtM2jwUg?e=Dkt7Vd

## Acknowledgments

We extend a word of thanks to King Abdullah International Medical Research Center (KAIMRK) for providing us with the data. In addition, our deepest gratitude goes to Dr. Khalid AlJarrah, Dr. Anwar Borai, Dr. Tariq Karrar, Dr. Muhammad Abdulfattah, and Dr. Jahangir Iqbal for providing us with useful research directions.

## Data availability statement

The data is available from the corresponding author on reasonable request.

