## Supplementary material for "Studies on Geographic Variations and Gender Bias in Thyroid Cancer at National Guard Hospitals in Saudi Arabia": https://1drv.ms/f/s!Ai2Oaa5jGDDxgZQtKjFKpvWtM2jwUg?e=Dkt7Vd

|  | Female | Male |
| --- | --- | --- |
| Anaplastic thyroid carcinoma | 13 | 10 |
| Follicular thyroid carcinoma | 70 | 16 |
| Hurthle cell thyroid carcinoma | 15 | 2 |
| Medullary thyroid carcinoma | 12 | 13 |
| Papillary thyroid carcinoma | 1459 | 350 |
| Thyroid lymphoma | 2 | 0 |
| Unspecified type of thyroid cancer | 159 | 45 |
