## Supplementary figures and images for "Studies on Geographic Variations and Gender Bias in Thyroid Cancer at National Guard Hospitals in Saudi Arabia"

### https://1drv.ms/f/s!Ai2Oaa5jGDDxgZQtKjFKpvWtM2jwUg?e=Dkt7Vd

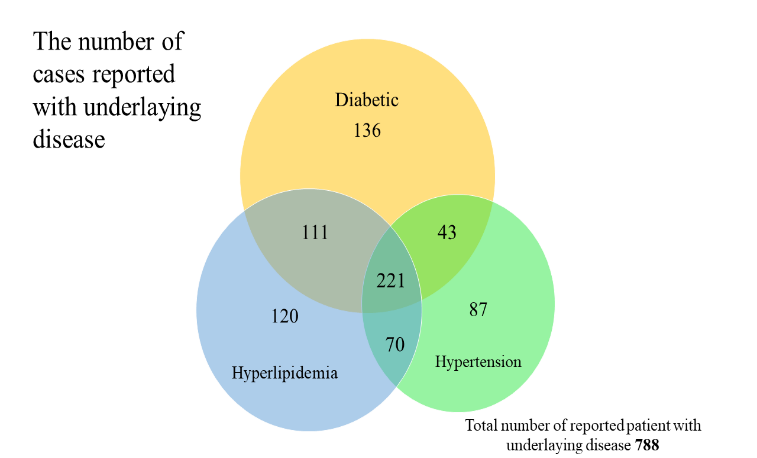

### https://1drv.ms/f/s!Ai2Oaa5jGDDxgZQtKjFKpvWtM2jwUg?e=Dkt7Vd

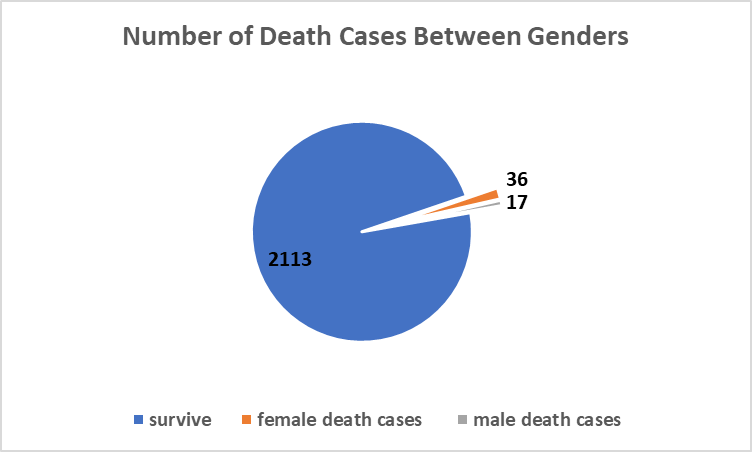
